# To what extent does self-reported physical activity impact children’s wellbeing and mental health? Insights from School-Aged Children in Wales

**DOI:** 10.1101/2024.11.06.24316822

**Authors:** M. James, M. Adebayo, M. Silveira Bianchim, L. Hughes, M. Mannello, E. Marchant, S. Brophy

## Abstract

There has been emphasis on the wellbeing of school-aged children amongst research, policy and practice in recent years, particularly during key developmental stages such as early childhood and adolescence. This study aimed to identify specific factors of self-reported physical activity that are associated with wellbeing and mental health among school-aged children in Wales. Using data from the Health and Attainment of Pupils in Primary Education in Wales (HAPPEN-Wales) cohort, this study analysed survey responses from 16,731 children aged 7-11 collected between 2016 and 2022. The HAPPEN survey includes self-reported measures of physical activity, physical literacy, sedentary behaviour, wellbeing, mental health, and local community environment. Wellbeing and mental health were assessed using the Good Childhood Index and the Me and My Feelings Questionnaire. Physical literacy was captured through questions on motivation, confidence, competence, and understanding of physical activity benefits. Additional data on environmental factors and socioeconomic status were also considered.

Multiple regression and decision tree analyses were used to examine factors associated with overall wellbeing. Key factors associated with good wellbeing included being more physically active (Coef.:0.17 [95%CI: 0.05 to 0.29]), being less sedentary (Coef.: -0.16 [95%CI: -0.28 to -0.05]), feeling confident to take part in lots of physical activity (Coef.: 0.35 [95%CI: 0.16 to 0.55]), feeling good at lots of physical activity (Coef.: 0.30 [95%CI:0.14 to 0.46], having less knowledge around understanding (Coef.: -0.29 [95%CI: -0.57 to -0.07]), feeling safe (Coef.: 0.77 [95%CI: 0.66 to 0.89]), autonomous (Coef.: 0.57 [95%CI: 0.40 to 0.73]) and competent (Coef.: 0.51 [95%CI: 0.35 to 0.67]).

The findings highlight the importance of providing opportunities for children to develop their confidence, competency and to feel like they have a choice in their lives. This can be done by providing safe, engaging, and varied physical activity opportunities in schools and communities to support children’s overall wellbeing.

## Introduction

Wellbeing is defined as “the state of feeling healthy and happy” (1). It is a multifaceted concept, typically encompassing positive psychological experiences (2), including mental health, happiness and life satisfaction (3), as well as physical and environmental factors (2) such as activity and opportunities. In recent years, there has been a focus in the United Kingdom (UK) policy landscape on the wellbeing of school-aged children (4). Early life, adolescence and early adulthood have all been marked as key time points in which wellbeing can be influenced (4,5). Evidence suggests that play and physical activity could be a key promoter of wellbeing and good mental health in young people. This was highlighted during the pandemic where school closures and transmission measures reduced access for children to be able to play and socialise (6,7).

Understanding the relationship between wellbeing, mental health and physical activity, particularly at an early age, could help inform the design of policy and practice aiming to develop and protect opportunities for movement, play, activity, and education for young people. Physical literacy [PL] is thought to lay the foundations of engagement with sport and other physical activities (8). It is defined as “*the motivation, confidence, physical competence, knowledge and understanding to value and take responsibility for engagement in physical activities for life*” (9). PL has a role in developing core skills that not only relate to being physically active but also core characteristics of good wellbeing such as confidence (10). Higher levels of PL likely lead to a lifelong engagement with activity (11–14) and thus, better wellbeing outcomes due to the positive associations of physical and mental health with increased physical activity.

School are a key setting in which wellbeing and mental health can be impacted. In 2022, Wales began the rollout of a reformed national curriculum, moving away from “narrow” subject areas and a prescriptive curriculum to broader, more holistic Areas of Learning Experience and school-level curriculum design (15). For example, schools can make strong connections between the Health and Wellbeing AoLE and the Literacy, Languages and Communication AoLE to address concerns regarding physical activity and diet (16). Within the CfW it is recommended to provide 120 minutes of PE a week, though this is not statutory (17). This is the same guidance given across the four nations however, research in 2018 found that 69% of schools met this (18). A recent report from Play Wales also noted a decline in self-directed, outdoor play (18). (19). The Active Healthy Kids [AHK] Report Card states that less than half of schools (45%) in Wales offer an afternoon break (20). These observations paint a concerning picture for children and young people’s opportunities to be active within the school day, this is essential to consider given its proposed relationship with wellbeing, mental health, learning and attainment.

Given the well-reported links between physical activity, wellbeing and mental health, this study set out to answer the question: *to what extent does self-reported physical activity impact wellbeing and mental health in children aged 7-11*? Our hypothesis assumes that physical activity and all attributed opportunities (i.e., active travel, riding a bike, feeling safe, perceptions of physical literacy) will have a significant impact on general wellbeing given existing research. This study uses data from Health and Attainment of Pupils in Primary EducatioN-Wales [HAPPEN-Wales] (21), a pan-Wales primary school cohort established by Swansea University in 2015 to explore the physical, social, emotional and mental health of children aged 7-11.

## Materials and Methods

HAPPEN-Wales (21) was established in 2014 by researchers at Swansea University and informed by research with head teachers from local primary schools (22,23). It was co-produced to provide a more collaborative and cohesive approach to health and wellbeing in schools, bringing together schools, partners in health and research to make more targeted health and wellbeing plans based on individual school needs as voiced by their pupils (22,24). HAPPEN has worked closely with school staff and pupils to develop the HAPPEN Survey; a self-report survey which is completed by primary school-aged children [aged 8–11, with expansion later to 7 years old] in education settings. The HAPPEN Survey captures a range of self-reported health behaviours including physical activity and sedentary behaviour, physical literacy, diet and dental health, wellbeing and mental health and the local community indicators. Schools receive an individual report of pupils’ group-level health and wellbeing data compared to national averages enabling tailored curriculum design for their Curriculum for Wales (CfW) and the health and wellbeing Area of Learning Experience (AoLE).

Since September 2014, over 600 schools have been invited to share details of the survey [including study aims and a parent information sheet] with parents/guardians so that parents can opt their child out of the survey (21). This opt-out method of recruiting participants was introduced in 2019 following the national roll-out in 2018, prior to this stage the network has been conducting more local and regional work in and around Swansea. This is informed via parent information sheets which are sent out prior to survey completion. All pupils in years 4, 5 and 6 can take part in the survey which is available in English and Welsh since September 2014 and is ongoing. The survey was co-developed alongside teachers, pupils, public health, and local authority teams. HAPPEN now includes a cohort of over 40,000 pupils across Wales. This aimed to ensure that a representative sample was recruited to reflect all children in Wales. The survey can be seen as supplementary file 1.

### Ethics Statement

This study was conducted with the approval of Swansea University’s Medical School Ethics Board (#7933). HAPPEN has had approval from the board since the project’s inception in 2014 and ethical approval was in place for the data collection period studied in this paper (2017–0033). Prior to 2019, parents and children provided written consent via consent forms. In 2019, this was amended to opt-out consent to overcome bias in the cohort. Parents can opt their child out of completing the survey by filling in the opt-out form which is received by the HAPPEN research team, the school is then notified and a record of this written opt-out is kept for records. Written consent from children is obtained at the start of the survey. This is informed via children’s information sheets (which have been informed by teachers, parents and children) which are sent out prior to survey completion and are at the beginning of the survey. Prior to starting the survey, children must consent to having their data used for research purposes. If a child does not consent but chooses to complete the survey, their record is removed. This process has been approved by Swansea University’s Medical School Ethics Board.

### Data Collection

The study uses data from 12^th^ September 2017 to 23^rd^ June 2023. This time frame encapsulates the national roll-out and six full academic years to provide a representative sample over a period.

This study aimed to ascertain the extent to which self-reported physical activity impacts wellbeing and mental health. The general wellbeing of children was measured by the validated Good Childhood Index [GCI] (25). The Children’s Society note that it is a statistically robust measure of the main aspects of children’s lives. Questions ask children to score their happiness with their a) friends, b) family, c) school, d) health, and e) life out of 10 (low happiness=1). For each component of the GCI, scores >=8 were assigned a 1 and <=7 a 0. To answer our question, we equated a total wellbeing score from this and a cumulative number was assigned based off the 5 GCI scores [highest score=5].

Mental health was measured by the Me and My Feelings Questionnaire [MMF] (26). The validated MMF measure captures emotional and behavioural mental health difficulties. In this study, both domains were utilised to assess mental health. Responses to the 16-item MMF are scored as 0, 1, or 2, corresponding to ‘Never’, ‘Sometimes’, and ‘Always’ respectively. These scores are then aggregated to derive an overall score. For the Emotional Difficulties Subscale, scores of 10 and 11 indicate borderline difficulties, and scores of 12 and above indicate clinically significant difficulties; for the Behavioural Difficulties Subscale, scores of 6 indicate borderline difficulties, and scores of 7 and above indicate clinically significant difficulties. These thresholds have been defined by Deighton et al. (2013) (26) and are valid and reliable in identifying clinically significant difficulties. Further to this, a binary outcome was established where a score of 1 represents clinical scores and 0 represents scores below the aforementioned thresholds.

Physical activity and environmental survey items were included in the analysis. This included the ability to ride a bike without stabilisers [yes or no], the ability to swim 25 metres without armbands [yes or no], use an active travel method to and from school [yes or no], physical activity (getting out of breath) for 60 minutes every day in the previous week [yes or no], two hours of sedentary time every day in the previous week [yes or no], safety of local area [high or low], ability to make your own decisions [yes or no] and feeling confident you are doing well [yes or no]. Deprivation scores of the child’s home were obtained from the Welsh Index of Multiple Deprivation (WIMD) (27). WIMD is based on numerous indicators such as income, access to services, safety, housing, and education. Quintiles were used in analysis with 1 equating to the most deprived area and 5 to the least deprived. Self-reported physical literacy data was collected in HAPPEN via 4 questions with a Likert response [strongly agree, agree, disagree, strongly disagree]; *a] I want to take part in physical activity, b] I feel confident to take part in lots of different physical activities, c] I am good at lots of different physical activities, and d] I understand why taking part in physical activity is good for me.* For this study, a cumulative score for physical literacy was obtained by assigning a binary outcome of 1 to responses of agree and strongly agree and a 0 to responses of disagree and strongly disagree. The cumulative number of 1 response was combined to generate a PL score [highest score=4].

### Analysis

Prior to beginning analysis, all data was anonymised, cleaned and coded by MJ. Data was accessed from 04/09/2023 for analysis. A linear regression model was used to explore the relationship between physical activity and components of physical activity (including environmental) on general wellbeing in the first instance. A decision tree was used to visually represent this relationship. The research team then used logistical models to explore the relationship between physical activity and mental health. Once both models were produced, variables were removed in a stepwise manner, removing potential explanatory variables in order and testing for significance after each step. As well as this, a decision tree analysis was also conducted. This dual approach aimed to provide a comprehensive understanding of how physical activity impacts wellbeing.

Analysis was undertaken by three researchers between September 2023 and June 2024. AM devised the analysis plan and initially completed this in R. Researchers MSB and MJ replicated the analysis in SPSS. This was to ensure the reproducibility, reliability, and validity of the findings. A breakdown of variables used in the analysis can be seen as supplementary file 2.

## Results

This study included a total of 16,731 participants, divided almost equally between boys (46.5%) and girls (49.2%), with a small percentage (4.2%) not specifying their gender. Participant characteristics are provided in Table 1. A significant proportion of the study population was from the most deprived quintiles, with 33.2% of boys and 34.1% of girls in the most deprived category, and a smaller representation from the least deprived quintiles. Most participants reported high overall wellbeing (the cumulative score of the GCI), with 49.6% of boys and 50.4% of girls in the high category. They also mostly reported good mental health with low numbers emotional and behavioural difficulties reported. Although girls reported higher emotional and boys reported higher behavioural.

**Table 1.**
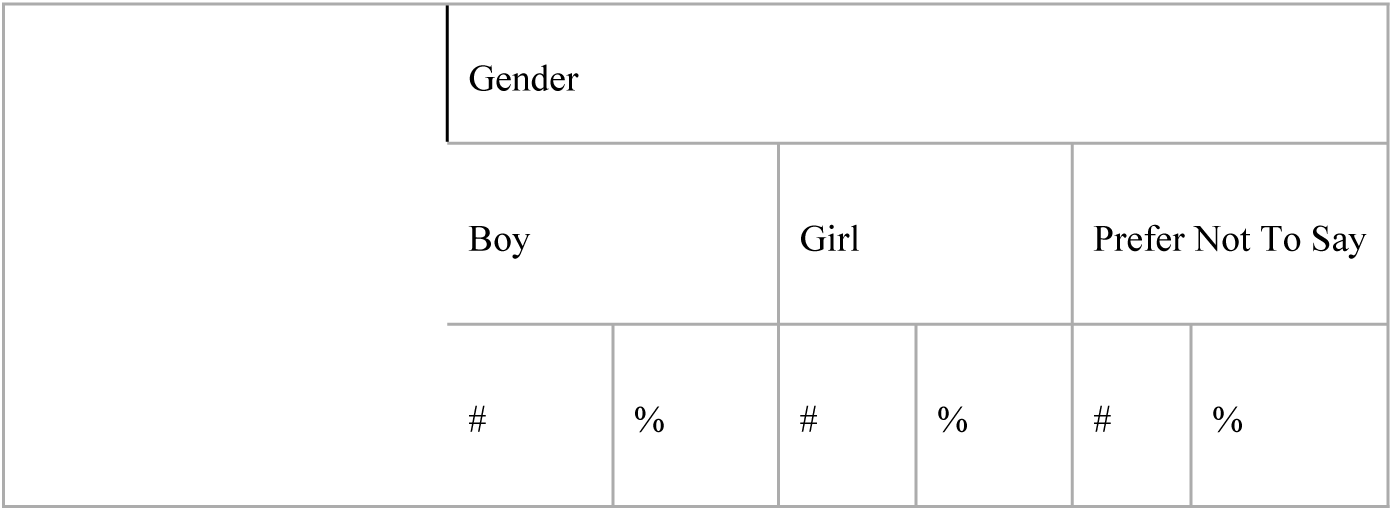
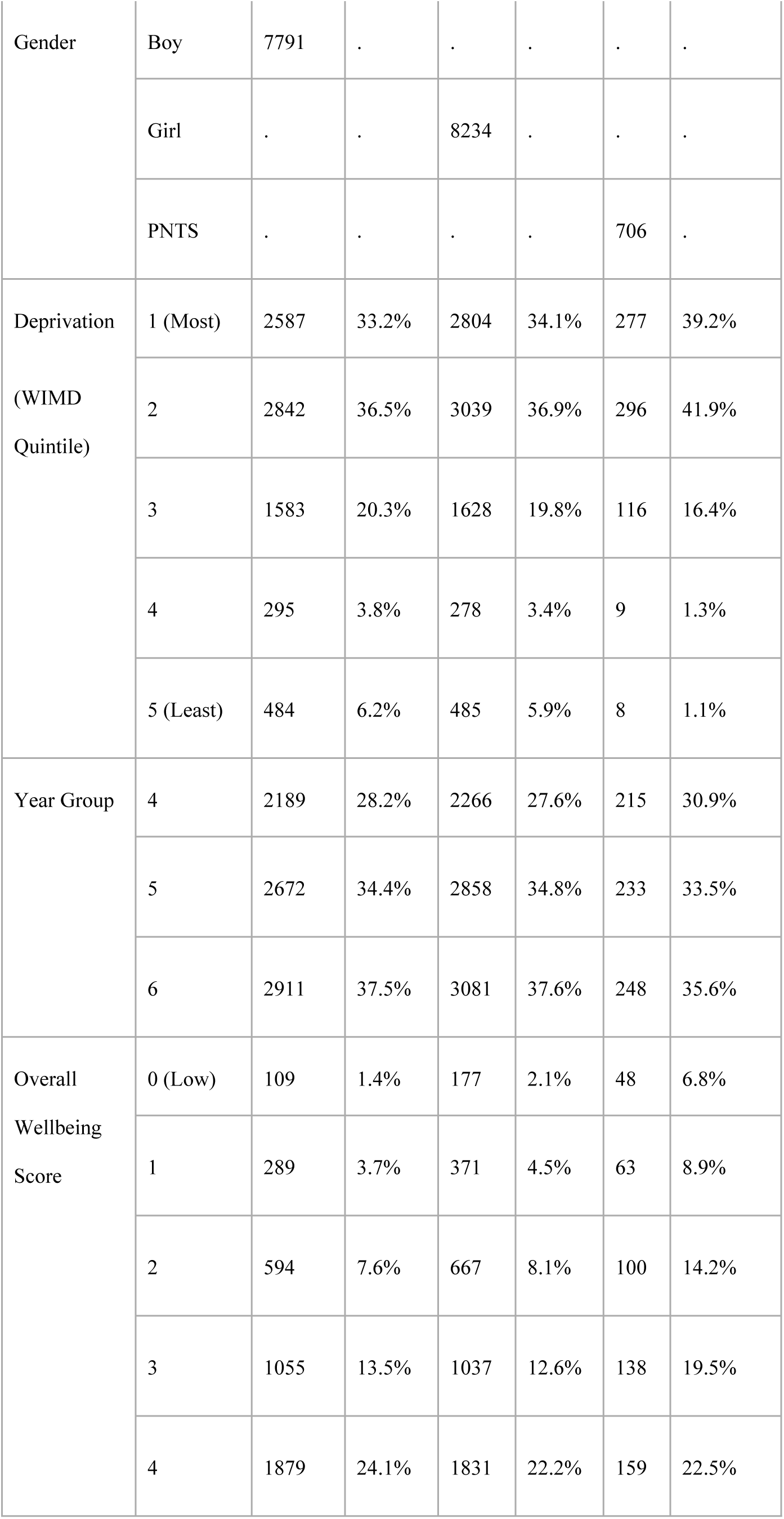
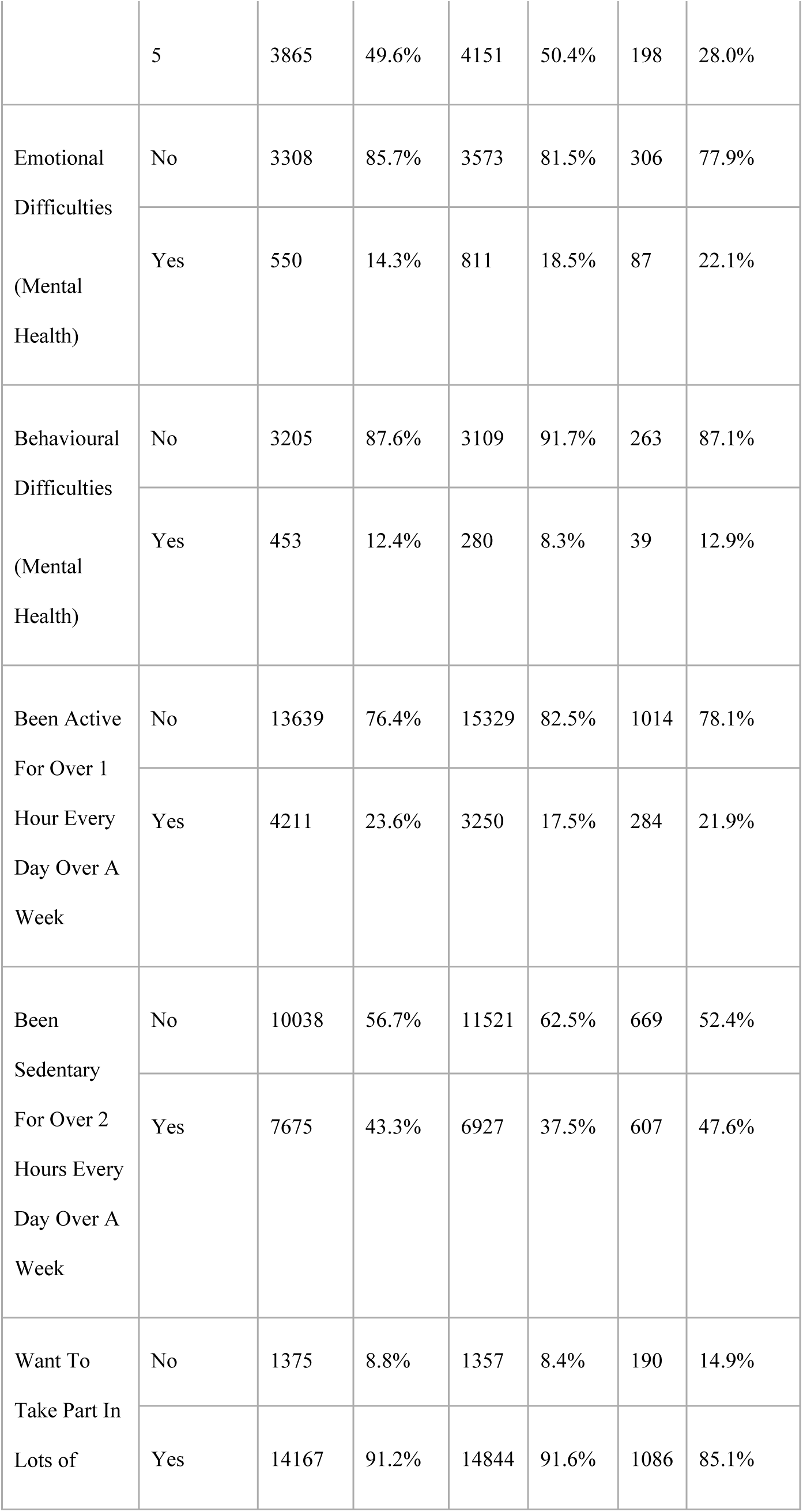
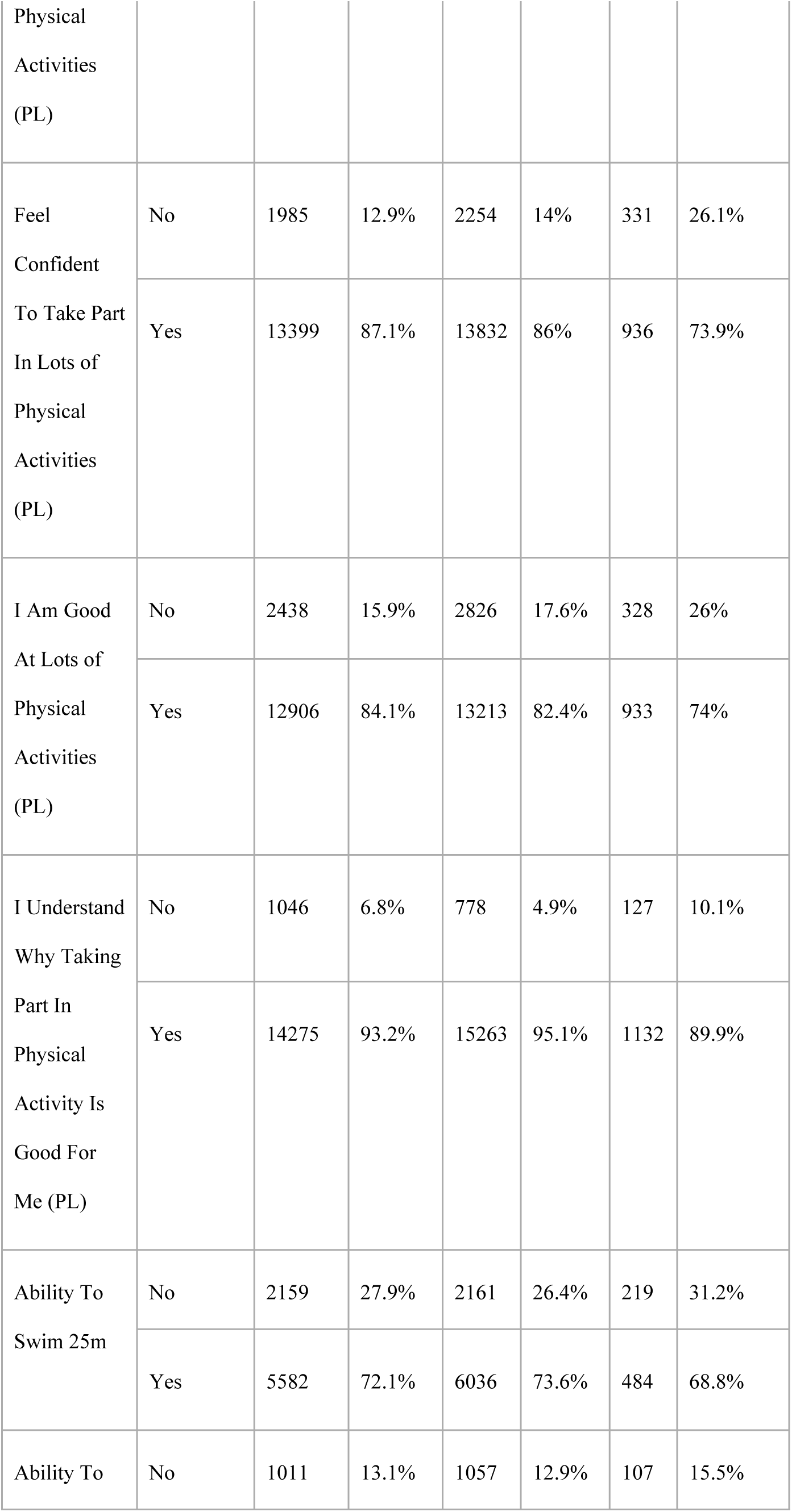
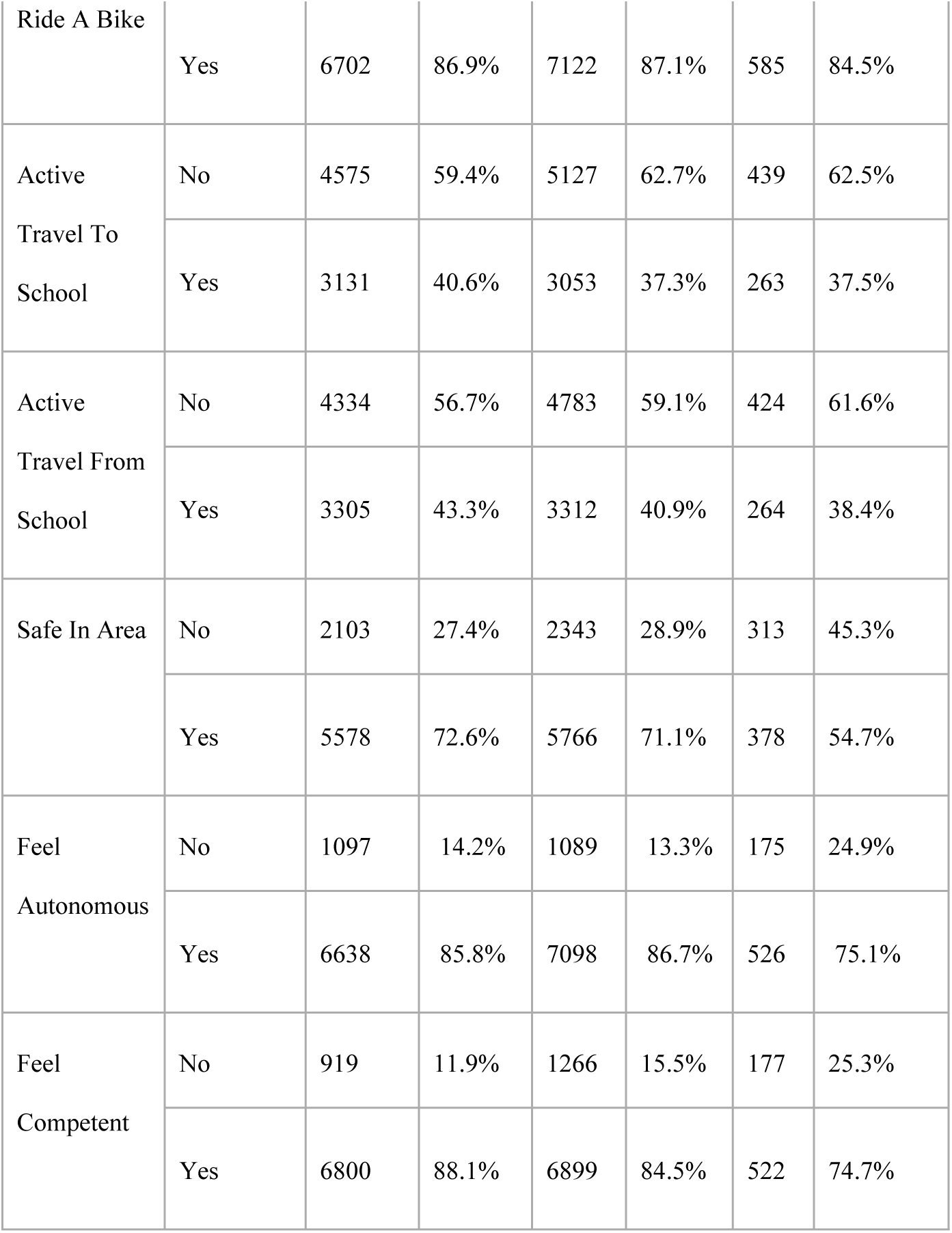
Characteristics of participants (n=16,731)

### Relationship Between Physical Activity and Overall Wellbeing

The regression model (table 2) indicated there are a number of predictors of good general wellbeing. Specifically, this included being more physically active (Coef.: 0.17 [95%CI: 0.05 to 0.29]), being less sedentary (Coef.: -0.16 [95%CI: -0.28 to -0.05]), feeling confident to take part in lots of physical activity (Coef.: 0.35 [95%CI: 0.16 to 0.55]), feeling good at lots of physical activity (Coef.: 0.30 [95%CI: 0.14 to 0.46], having less knowledge around why physical activity is good for you (Coef.: -0.29 [95%CI: -0.57 to -0.07]), feeling safe in your area (Coef.: 0.77 [95%CI: 0.66 to 0.89]), autonomous (Coef.: 0.57 [95%CI:0.40 to 0.73]) and competent (Coef.: 0.51 [95%CI: 0.35 to 0.67]).

**Table 2.**
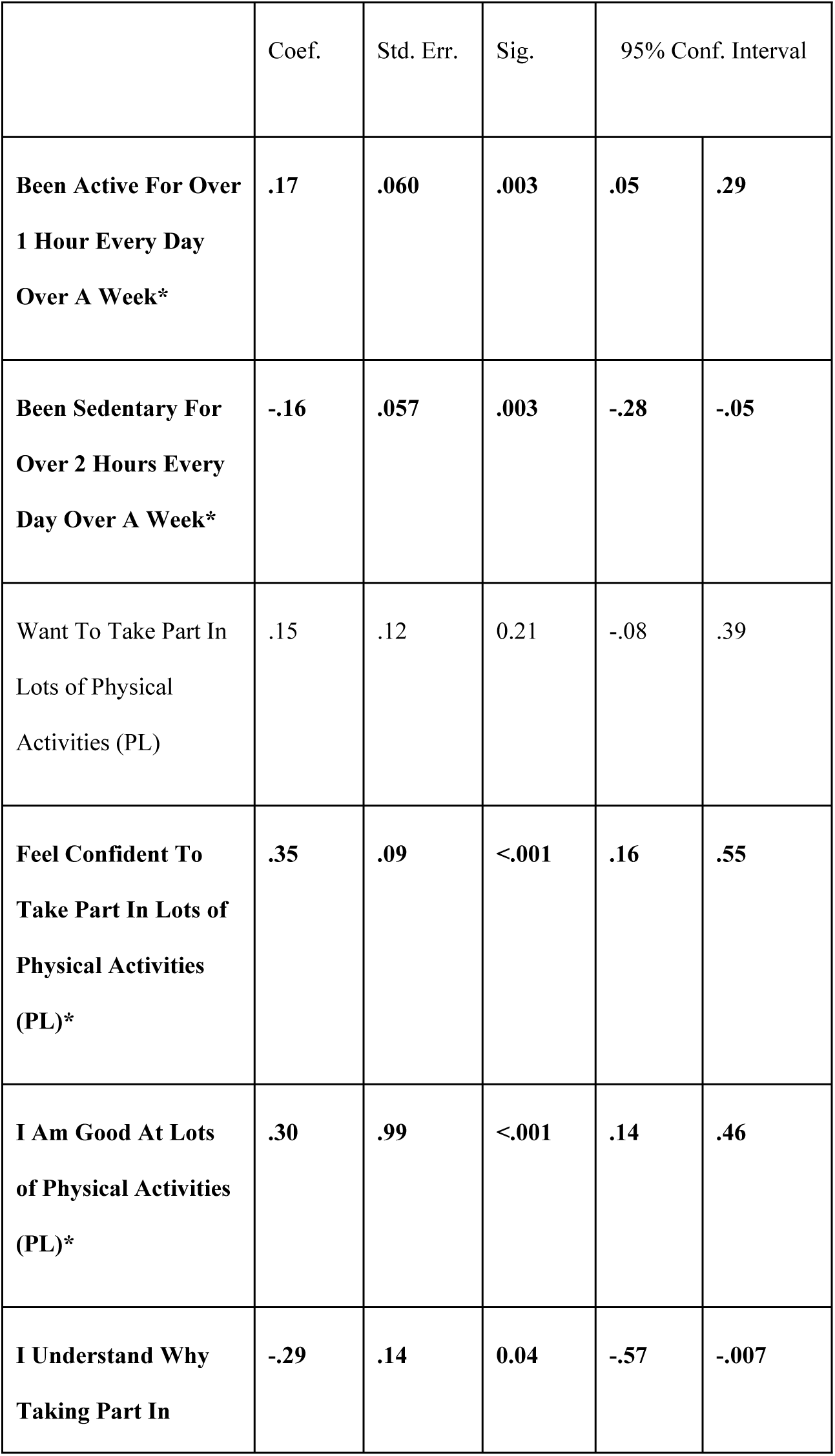
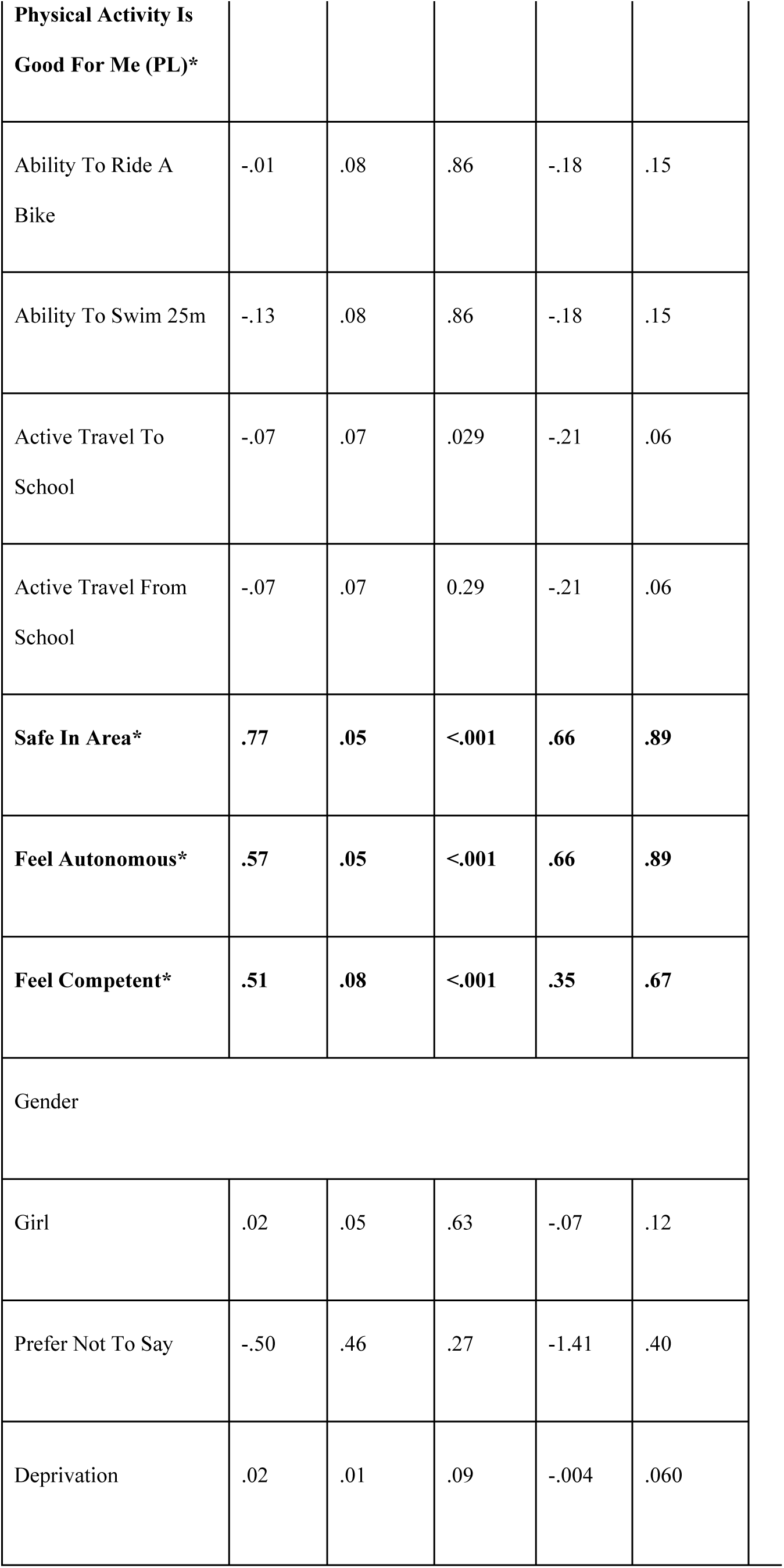
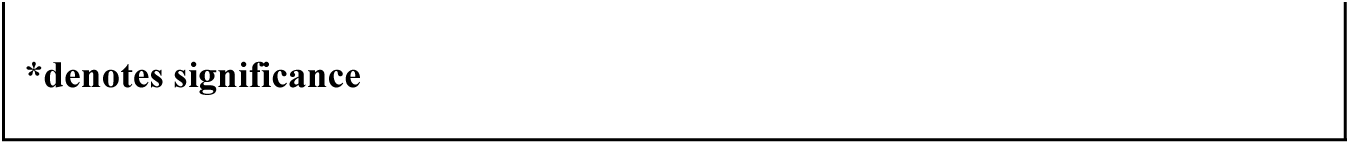
Activity based predictors of general wellbeing.

Interestingly, gender and deprivation had no significant role to play in predicting general wellbeing for this age group. When removed in a stepwise manner, the most significant predictors were being less sedentary, feeling confident, feeling good, swimming (not being able to swim 25m), feeling safe, feeling autonomous and feeling competent. Thus, suggesting that positive intrinsic feelings around being active are more important than specific indicators of physical activity participation.

### Relationship Between Self-Reported Physical Activity and Mental Health

Table 3 shows key predictors of lower emotional wellbeing (indicated by presenting clinical levels of emotional difficulties based on the cut-offs provided by the Me and My Feeling Questionnaire) included being more sedentary (Coef: 0.35 [95%CI: 0.09 to 0.61]), feeling less confident (Coef:. -0.60 [95%CI: - 0.99 to -0.20]), feeling less safe in your area (Coef.: -0.99 [95%CI: -1.25 to -0.73), feeling less competent (Coef.: -0.58 [95%CI: -0.92 to -0.25]) and being a girl (Coef.: 0.49 [95%CI: 0.24 to 0.74]).

**Table 3.**
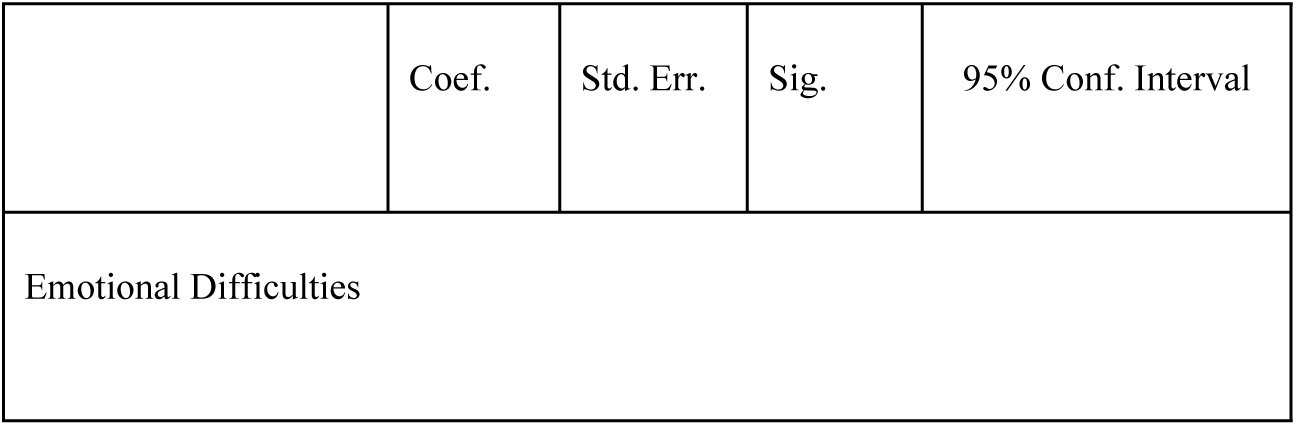
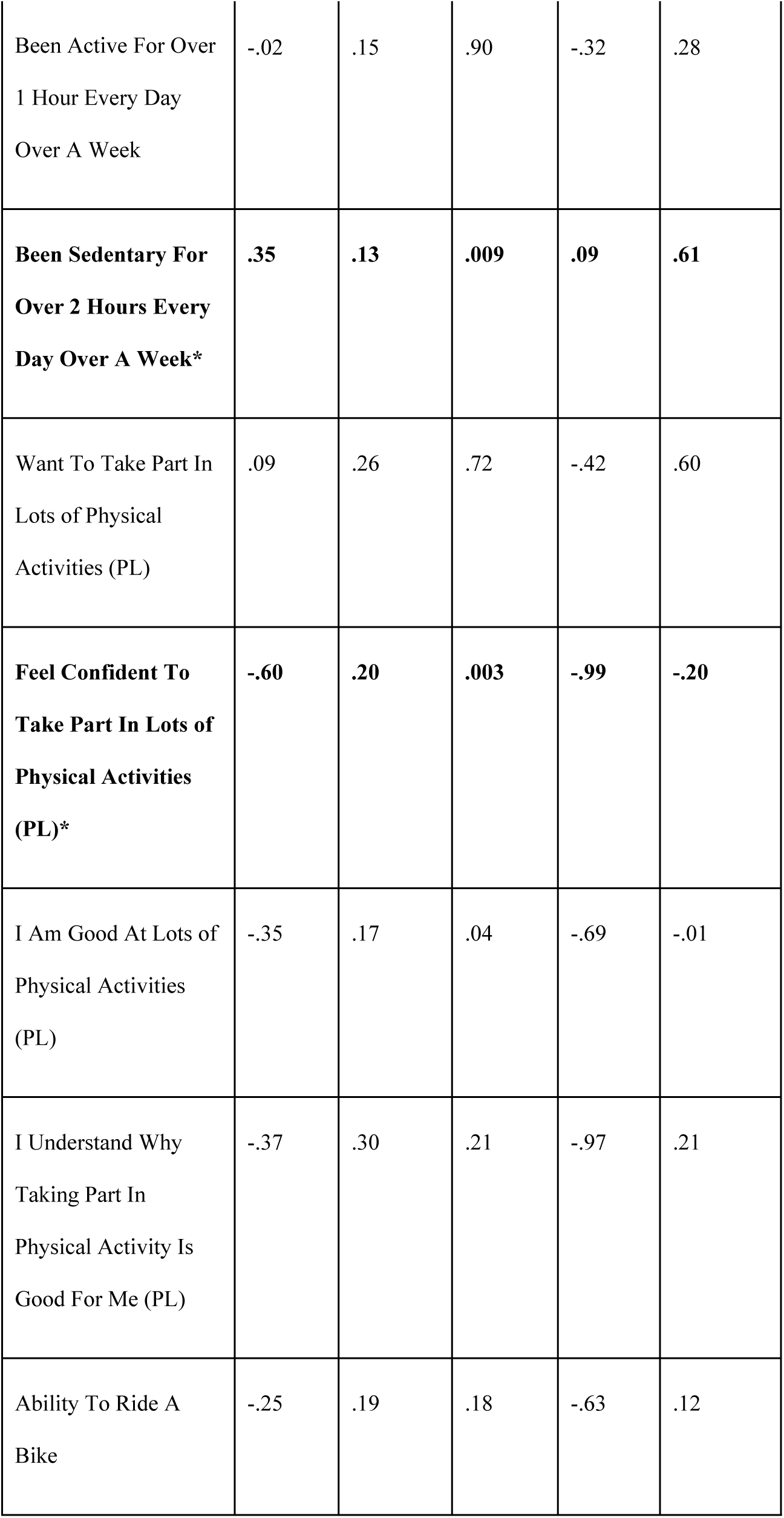
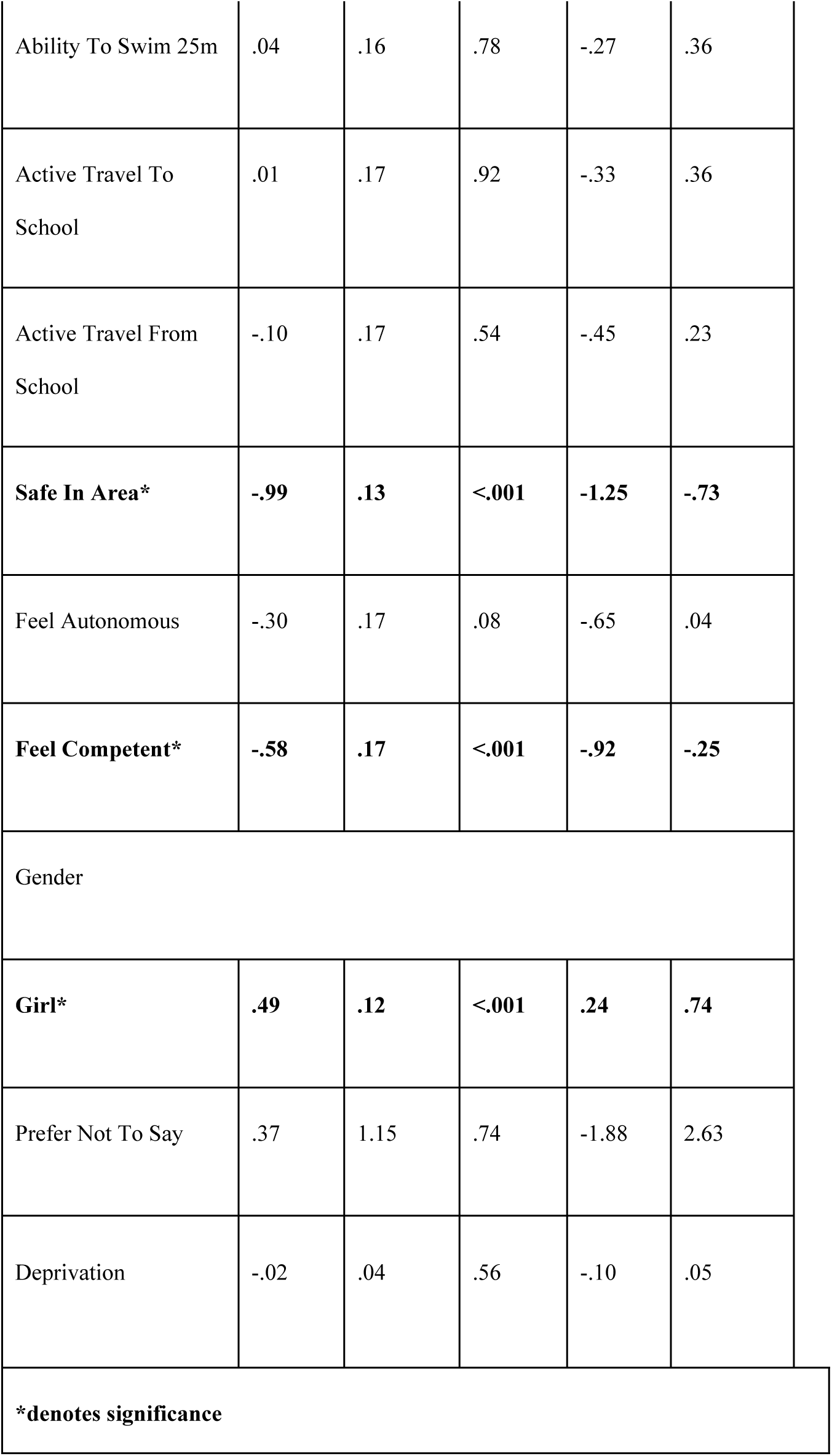
Self-Reported Physical Activity-based predictors of emotional difficulties.

|  | Coef. | Std. Err. | Sig. | 95% Conf. Interval |
| --- | --- | --- | --- | --- |
| Emotional Difficulties |  |  |  |  |

|  |  |  |  |  |  |
| --- | --- | --- | --- | --- | --- |
| Been Active For Over<br>1 Hour Every Day<br>Over A Week | -.02 | .15 | .90 | -.32 | .28 |
| **Been Sedentary For**<br>**Over 2 Hours Every**<br>**Day Over A Week\*** | **.35** | **.13** | **.009** | **.09** | **.61** |
| Want To Take Part In<br>Lots of Physical<br>Activities (PL) | .09 | .26 | .72 | -.42 | .60 |
| **Feel Confident To**<br>**Take Part In Lots of**<br>**Physical Activities**<br>**(PL)\*** | **-.60** | **.20** | **.003** | **-.99** | **-.20** |
| I Am Good At Lots of<br>Physical Activities<br>(PL) | -.35 | .17 | .04 | -.69 | -.01 |
| I Understand Why<br>Taking Part In<br>Physical Activity Is<br>Good For Me (PL) | -.37 | .30 | .21 | -.97 | .21 |
| Ability To Ride A<br>Bike | -.25 | .19 | .18 | -.63 | .12 |
| Ability To Swim 25m | .04 | .16 | .78 | -.27 | .36 |
| Active Travel To School | .01 | .17 | .92 | -.33 | .36 |
| Active Travel From School | -.10 | .17 | .54 | -.45 | .23 |
| **Safe In Area\*** | **-.99** | **.13** | **<.001** | **-1.25** | **-.73** |
| Feel Autonomous | -.30 | .17 | .08 | -.65 | .04 |
| **Feel Competent\*** | **-.58** | **.17** | **<.001** | **-.92** | **-.25** |
| Gender |  |  |  |  |  |
| **Girl\*** | **.49** | **.12** | **<.001** | **.24** | **.74** |
| Prefer Not To Say | .37 | 1.15 | .74 | -1.88 | 2.63 |
| Deprivation | -.02 | .04 | .56 | -.10 | .05 |
| **\*denotes significance** |  |  |  |  |  |

The stepwise removal of variables showed that confidence, safety, competence and gender are significant predictors of emotional difficulties and therefore, poorer mental health in this age group. These findings show that intrinsic motivators are of more value than specific indicators of physical activity participation and levels.

A similar picture is observed with behavioural difficulties (table 4) with being more sedentary (Coef.: 0.31 [95%CI: 0.15 to 0.47]), feeling less safe (Coef.: -0.44 [95%CI: -0.60 to -0.27]), feeling less autonomous (-0.26 [95%CI: -0.47 to -0.05]), feeling less competent (Coef.: -0.31 [95%CI: -0.53 to -0.09]) also showing trends toward poorer mental health outcomes. Interestingly, for this measure of being more confident (Coef.: 0.31 [95%CI: 0.15 to 0.47]) was associated with less behavioural problems and being a boy (Coef.: -0.45 [95%CI: -0.62 to -0.28]) was associated with more behavioural problems. This was the first model in which deprivation was significant, lower deprivation was associated with a lower trend of having a behavioural difficulty (Coef:. -0.12 (95%CI: -0.19 to -0.05]).

**Table 4.** Activity based predictors of behavioural difficulties.

|  | Coef. | Std. Err. | Sig. | 95% Conf. Interval |  |
| --- | --- | --- | --- | --- | --- |
| Behavioural Difficulties |  |  |  |  |  |
| Been Active For Over<br>1 Hour Every Day<br>Over A Week* | .07 | .10 | .45 | -.12 | .27 |
| **Been Sedentary For**<br>**Over 2 Hours Every**<br>**Day Over A Week\*** | **.31** | **.08** | **<.001** | **.15** | **.47** |
| Want To Take Part In Lots of Physical Activities (PL) | -.08 | .15 | .59 | -.38 | .22 |
| **Feel Confident To Take Part In Lots of Physical Activities (PL)\*** | **.03** | **.13** | **.77** | **-.22** | **.29** |
| I Am Good At Lots of Physical Activities (PL) | -.002 | .11 | .98 | -.23 | .22 |
| I Understand Why Taking Part In Physical Activity Is Good For Me (PL) | -.15 | .16 | .37 | -.48 | .17 |
| Ability To Ride A Bike | .06 | .12 | .59 | -.17 | .30 |
| Ability To Swim 25m | .04 | .09 | .63 | -.14 | .23 |
| Active Travel To School | .13 | .12 | .11 | -.04 | .43 |
| Active Travel From School | -.24 | .12 | .04 | -.47 | -.27 |
| **Safe In Area\*** | **-.44** | **.08** | **<.001** | **-.60** | **-.27** |
| **Feel Autonomous\*** | **-.26** | **.10** | **.01** | **-.47** | **-.05** |
| **Feel Competent\*** | **-.31** | **.11** | **.005** | **-.53** | **-.09** |
| Gender |  |  |  |  |  |
| **Girl\*** | **-.45** | **.08** | **<.001** | **-.62** | **-.28** |
| Prefer Not To Say | -.25 | .19 | .19 | -.64 | .12 |
| **Deprivation\*** | **-.12** | **.03** | **<.001** | **-.19** | **-.05** |
| **\*denotes significance** |  |  |  |  |  |

Stepwise removal showed that higher sedentary time, less autonomy and gender (being a boy) were key predictors for high behavioural difficulties. This is the first wellbeing/mental health indicator in which physical activity participation is not associated with the outcome (e.g. PA does not improve or worsen behavioural difficulties).

### Decision tree analysis

The decision tree analysis in Figure 1 investigates the clustering of factors predicting overall wellbeing among school children. The most important predictor of wellbeing is a sense of competence. Then among those who feel competent, autonomy is the next most important variable followed by participation in physical activities, particularly for those who also feel autonomous.. However, low emotional wellbeing is an important negative factor that impacts overall wellbeing, especially for those who do not feel competent or autonomous. For those lacking competence or autonomy then confidence in physical activities can improve wellbeing.

**Figure 1.**
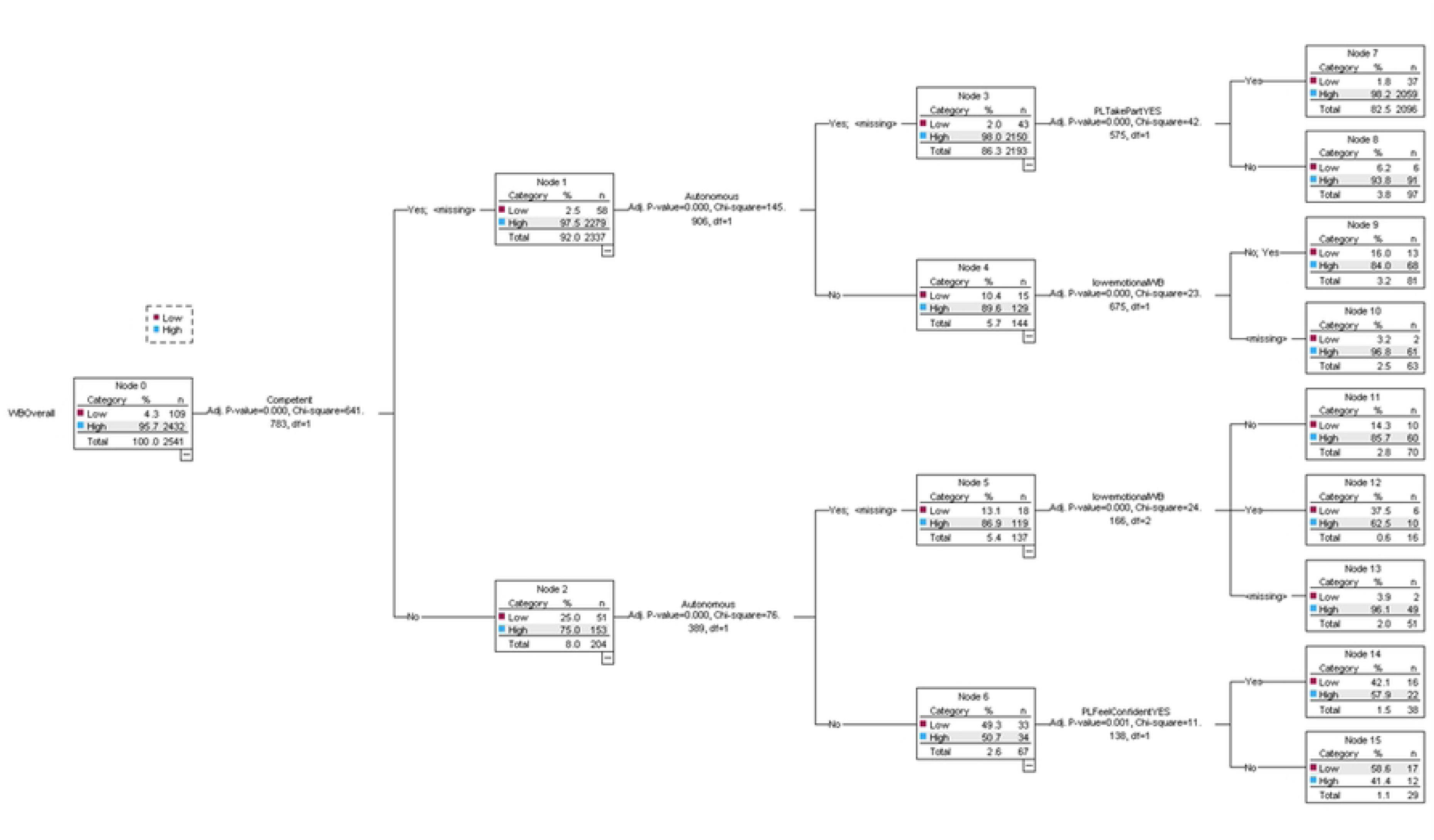
Decision tree analysis.

## Discussion

This study explores the key physical activity predictors of wellbeing and mental health in school-aged children in Wales using HAPPEN-Wales. Findings aim to provide evidence that would help schools, communities and policy makers understand how physical activity provision should be designed and tailored with this in mind. The links between physical activity and wellbeing and mental health are well-established. It is well-documented that regular physical activity enhances mood, reduces anxiety and depression and improves wellbeing (3). However, it remains unclear which attributed characteristics (i.e., active travel, riding a bike, feeling safe, perceptions of physical literacy) have a significant impact on general wellbeing.

Overall, analysis revealed that key predictors of good wellbeing and mental health are low sedentary time, high confidence, high autonomy, high safety and male gender. This is a key finding as it could help inform tailored interventions for young people to help reduce inequalities in wellbeing. Interestingly, this study highlights that intrinsic feelings around activity participation which are integral and that opportunities which protect and enhance confidence, autonomy and safety for girls and boys from a variety of backgrounds could be the solution to improve physical activity and, in turn, creating healthier and happier children and young people.

### Overall Wellbeing

The most significant indicators of good wellbeing were being less sedentary, feeling confident, feeling good, not doing swimming lessons (not being able to swim 25m), feeling safe, feeling autonomous and feeling competent. While self-reported physical activity time was positively associated with wellbeing, there were also a variety of intrinsic factors at play, showing that increasing the time to be active alone is not necessarily the solution needed. While regular physical activity in childhood is more likely to lead to move physically active adolescents and adults (28), it is important that children and young people are provided with a variety of opportunities to be active in every setting (school, home, after school), this study shows that these opportunities should improve a child’s confidence. It is noteworthy that not being able to swim was associated with high wellbeing, advocating for less adult-led forms of activity provision. For example, research shows that walking can be an important activity (29,30), therefore employing ways in which this could be promoted would be significant. Evidence also reports positive impacts on children’s PA levels, social engagement and emotional wellbeing from unstructured play, this is play which is self-directed with children doing what they wish, in their own way, in their own time (31).

Play supports children in developing their mental, physical, social and emotional wellbeing (32). The Foundation Phase and its play-based methods are praised by stakeholders in Wales (33) for providing safe and engaging places to play. However, play opportunities have been declining, research suggests that reductions in play is due to the focus on core subjects (e.g., literacy and numeracy), a lack of adult confidence in play expertise and in turn, the perspective of teachers that play does not have a formal status within their practice (33). Children and young people have consistently asked for more active opportunities. In schools, children have asked for more time, space and permission to be active and play (30,34) advocating for more opportunities to have less formal learning and more time to engage in play in a variety of ways with their friends. This trend has been observed throughout the years but has been highlighted by the pandemic where opportunities to play were limited. In comparison to interventions such as the Daily Mile (35), outdoor play can also ensure that more opportunities for PA are introduced into the school day and beyond.

The intrinsic benefits of child-led play and physical activity opportunities cannot be overlooked. It is these characteristics of confidence, competence and autonomy which have shown to be integral to good wellbeing. An underlying motivation for being active is the belief in being able to participate. This study demonstrated that wanting to take part in the activity, feeling confident to take part, feeling good at lots of activities, having autonomy and feeling competent to take part were key influences. Confidence and competence are also key components of PL (9) which can have a significant impact on wellbeing by providing the foundations for increased participation in physical activity. Research from Melby et al. (2022) (36) highlights that helping children develop PL is more beneficial for wellbeing than just trying to improve levels of PA in isolation, shifting the focus away from the intensity of activities to the quality and enjoyment of encouraging children to move. This approach would help children want to take part, feel confident to take part, feel competent to take part, feel good and have autonomy over their activity if the focus was on fun rather than time and effort.

This is important to note, particularly in Wales, where greater autonomy has been placed on schools within the CfW to influence health and wellbeing through school-level curriculum design (11), therefore providing opportunities to implement this ethos into the day-to-day of school-aged children. Guidance within the new curriculum regarding PA is broad and as a result, there are concerns regarding children and young people’s opportunities to play and be active. Several PL-based programs are being implemented. In Wales, the Dragon Challenge (37) has been used to measure PL for assessment purposes, as well as for national surveillance. Programmes such as this can help to monitor the impacts of statutory focus on health and wellbeing and intervention effects.

Autonomy allows individuals to feel that they can make their own choices. Choice has often been cited as an opportunity to improve physical activity for this age group, with prescriptive forms often seen as barriers due to their constraints on rules, times and resources. It is crucial to continue promoting a wider range of activities available for all children to support their wellbeing. It is important to note the key influences and ways in which we can continue to promote a wider choice of activity provision for this group. Thus, pupils and educational professionals should be involved in the design of such interventions based upon the wants and needs of children, factoring in structured and unstructured PA (play) and the creation of safe spaces.

Feeling safe in an area was the strongest positive predictor of overall wellbeing. This underpins the critical role that safe opportunities play in enhancing wellbeing. Safety has been highlighted by several studies as influential in supporting physical activity and wellbeing in general (3,29,30). Community characteristics such as crime rates, traffic and littering can all increase safety concerns. Parental influence and socio-economic status are also influential in safety perceptions, with more deprived children at risk of lower community safety (objectively or subjectively) and ultimately, reduced health and wellbeing (3,38,39). While it is harder to externally control safety, it is worth noting that children who fall into this category are more likely to have lower wellbeing as demonstrated by this study’s findings.

### Physical activity and mental health

Confidence, safety, competence and gender are significant predictors of emotional difficulties and higher sedentary time, less autonomy and gender (being a boy) were key predictors for high behavioural difficulties. The increased accessibility and use of screens has been shown to negatively impact wellbeing by contributing to higher levels of sedentary behaviour (40,41), therefore any provision that mitigates this is important. In schools, interventions such as the Daily Mile have been implemented as a social physical activity initiative that encourages children and young people to walk or run outdoors with their friends for 15 minutes each day (42). However, whilst evidence suggests the benefits to children’s PA levels, challenges include interventions like The Daily Mile replacing existing PA opportunities such as PE lessons or afternoon break times (43). They also show evidence of improving PA but no evidence of the impact on wellbeing or longer-term follow-up impacts (43). Shifting the focus away from the intensity of activities to the quality and enjoyment of encouraging children to move (36) could be a significant progression step in helping children be healthier and happier as demonstrated by the key predictors observed in this study.

Gender showed negative associations with mental health, indicating that girls (and individuals from more deprived areas) experience higher emotional difficulties and boys experiencing higher behavioural difficulties. These disparities may be due to a variety of factors, including different access, gender-specific expectations, resources and opportunities for PA. With girls significantly more likely to drop out of activity in their early teens, it is important that we uphold positive associations with PA and PL from an early age. Listening to what they want and need from PA opportunities could go some way to preventing this. This is also important to consider when tailoring interventions.

The decision tree analysis further cemented the factors influencing overall wellbeing. Our findings showed that feeling competent and safe significantly reduced emotional difficulties. Similarly, these factors, along with autonomy, correlated with lower behavioural difficulties. These findings highlight the protective role of feeling capable, safe and autonomous for young people. These are important characteristics as capability and autonomy can be underpinned by PL and PA respectively.

### Limitations

This study and its findings are built on a cohort of Welsh school-aged children from 2015 to 2022. Therefore, it may not be appropriate to generalise findings, particularly those that relate to the Welsh context including specific legislation and the Curriculum for Wales. Furthermore, the sample is based on schools who chose to actively engage with HAPPEN. However, lessons can be learned from these policies. Future research should also explore a broader range of predictors and consider longitudinal studies to assess the long-term impact of interventions targeting PL and PA on overall wellbeing. The findings are cross sectional surveys and so it is not possible to give causal inference that being active leads to better wellbeing just that children who self-report lower sedentary levels report better wellbeing.

### Conclusion

Early years is a pivotal timepoint in which wellbeing can be influenced (4,5) and therefore, has important policy and practice implications. The key message is that improving time and amount of physical activity on its own is not enough for wellbeing and mental health benefits alone, belief in being able to participate and opportunities to do PA can enhance wellbeing by addressing confidence, competence and autonomy. These can also be tailored for specific demographic groups, particularly girls and those from deprived areas to help reduce inequalities. Higher wellbeing equates to better learning outcomes which is likely to impact academic attainment (4), therefore it serves school settings to provide for and listen to children and young people.

Given the findings of this study, there are key considerations which could be made to improve the wellbeing and mental health of young people:

1. Involve pupils, teachers and other key stakeholders in co-designing PA/PL interventions. This ensures that they are relevant and meet the needs of the children. This will go some way to giving young people autonomy over their activity and needs.
2. Provide a variety of PA opportunities within schools, such as PE lessons, break-time activities, and extracurricular clubs. This should include structured and unstructured play to cater to different interests and promote overall wellbeing.
3. Work with local authorities to improve community safety by addressing crime rates, traffic, and littering, which can significantly impact children’s sense of safety and, subsequently, their wellbeing and mental health.

Helping children to have the confidence to feel that they are good at an activity, feeling safe and having autonomy is good for wellbeing. It is worth noting that play and physical activity can help with giving a sense of confidence and competence, but it might not be the only solution. Future work should look to explore other activities such as creative outlets.

## Funding Statement

This study has been funded by the Centre for Population Health (CPH), ADR-Wales and Play Wales. The funders had no role in study design, data collection and analysis, decision to publish, or preparation of the manuscript.

## Data Availability

Data cannot be shared publicly to protect the anonymity of participants. Data are available from upon reasonable request for researchers who meet the criteria for access to confidential data.

## Acknowledgements

The research team would like to thank all teachers, schools and pupils who have taken part in HAPPEN since its inception in 2014 and for their support in co-developing and producing the survey, strategy and direction.

## Author Contributions

MJ led the conceptualisation, data curation, analysis, methodology, visualisation and writing (original draft and ongoing revisions) of this study. MA assisted with analysis and methodology. MSB supported analysis and writing (original draft and ongoing revisions). MM assisted with writing (ongoing revisions) and visualisation. LH supported writing ongoing revisions. EM assisted with writing (ongoing revisions) and has helped with data curation. SB supported writing (ongoing revisions) and ongoing data curation.

## Notes

### Competing Interest Statement

The authors have declared no competing interest.

### Author Declarations

This study was conducted with the approval of Swansea University’s Medical School Ethics Board (#7933).

